# A hybrid approach to scalable real-world data curation by machine learning and human experts

**DOI:** 10.1101/2023.03.06.23286770

**Authors:** Michael L. Waskom, Katherine Tan, Holly Wiberg, Aaron B. Cohen, Brett Wittmershaus, Will Shapiro

## Abstract

**Objective:** Machine learning has the potential to increase the scale of real-world data curated from electronic health records, but maintaining a high standard of data quality is important to avoid biasing downstream analyses. To increase scale without compromising quality, we propose a hybrid data curation methodology that employs both manual abstraction by clinical experts and automated extraction by machine learning models.

**Materials and Methods:** Our methodology makes the determination about when to employ manual abstraction using a confidence score associated with each model output. We describe a process for selecting confidence thresholds based on simulations validated against a reference-standard labeled dataset. To establish the fitness of our methodology for retrospective research, we apply it to a multi-variable cohort selection task on a large real-world oncology database.

**Results:** Only small amounts of manual abstraction are required for hybrid curation to achieve expert-level error rates. In fact, the hybrid methodology can even reduce error rates relative to manual abstraction in some cases. We further demonstrate that demographic characteristics of a research cohort defined using hybrid variables are comparable to one curated with conventional methods.

**Discussion:** Our methodology is general and makes few assumptions about the clinical variable or machine learning model. A key requirement is the availability of reference standard labels for calibrating the tradeoff between abstraction effort and data quality.

**Conclusion:** Incorporating machine learning into real-world data curation using hybrid methodology holds the promise to scale practicable cohort sizes while maintaining data fitness for research purposes.

## Introduction

Retrospective analysis of real-world data (RWD) generated during routine clinical care has the potential to address important scientific and policy-related questions.^1,2^ Relative to traditional clinical trials, RWD-based research can address larger patient cohorts that are more representative and studied for longer durations.^3,4^ RWD can be derived from a number of sources, including tumor registries, insurance claims, and electronic health records (EHR). In particular, EHR data offer a deep and longitudinal perspective that may not be available from other sources.

While some information is captured in structured EHR fields, structured data often have missing values, encode information at only a high level, and fail to capture important datapoints such as diagnosis dates or biomarker test results.^5^ The depth and completeness of EHR-derived RWD can therefore be substantially enhanced by including information from unstructured sources such as clinical notes or reports. To be useful for quantitative analysis, however, this information must first be curated into structured datapoints. The curation process can be challenging because medical records are often complex and nuanced.^6^ Errors caused by oversight (e.g., missing a biomarker test result) or miscoding (e.g., mistaking a second primary tumor for a distant metastasis) have the potential to bias downstream analyses.^7^

To produce datasets that are fit for use in retrospective analyses, the RWD curation process must be reliable and unbiased. Traditionally, this has been achieved through manual curation by highly-trained abstractors. Yet manual abstraction is effortful and time consuming, and finite abstraction resources can limit the practicable size of RWD datasets.^5^ Therefore, realizing RWD’s promise of larger cohort sizes requires a curation process that is both accurate and scalable.

Recent advances in natural language processing (NLP) using machine learning (ML) algorithms^8^ have the potential to help realize this promise. Algorithmic approaches can rapidly process massive volumes of unstructured data, surfacing relevant information with high reliability and scaling to arbitrarily large cohorts with minimal marginal cost. But substituting ML algorithms for human abstractors may increase the risk of errors when charts are complex or require clinical expertise to correctly interpret. Therefore, it is important to understand how ML can be used to scale RWD curation while maintaining a level of accuracy that is fit for use in research applications.

Here, we present a novel methodology that achieves these objectives. Our approach has three important components: we employ a “hybrid” process such that, within a given variable (e.g., date of diagnosis), some datapoints are generated using only ML, while others are determined through manual review by an expert human abstractor; we use the likelihood that the model is correct (i.e., the model’s “confidence”) to allocate manual abstraction effort; and we create a set of reference standard labels to understand the tradeoffs between accuracy and effort. This allows us to reduce the manual effort needed for data curation—and hence scale practicable cohort sizes—without compromising the accuracy of the process. In fact, we demonstrate that hybrid variables can have lower error rates than variables produced using only manual abstraction. Therefore, our approach balances the strengths of manual and algorithmic RWD curation to produce datasets with scale and quality that could not be achieved through either method alone.

## Methods

This work is divided into two sections. We first describe the process of hybrid RWD curation. We then demonstrate its performance when applied to a large dataset of oncology records.

### Hybrid extraction methodology

#### RWD variables

RWD variables comprise structured datapoints representing information derived from medical records. The information may be structured at point of entry (i.e., entered using a dropdown menu in an EHR system) or curated from information present in unstructured records (clinical notes, surgical reports, etc.). This work focuses on the curation of RWD from unstructured sources (Adamson, et al. in press, 2023).

RWD variables may contain a single datapoint for each patient (e.g., the date of the patient’s initial diagnosis with a particular cancer) or multiple datapoints (e.g., the result of every test performed for a specific tumor biomarker). We refer to “datapoint” as the finest level of granularity relevant for a particular variable; concretely, it corresponds to a row in the structured output. While this manuscript focuses on the generation of patient-level datapoints, the methodology is general.

Critical to the curation of high-quality RWD is a set of clinically-backed standardized policies and procedures for abstracting information from clinical documents. These standardized policies aim to optimize accuracy and consistency, reduce ambiguity, and specify the handling of known edge cases. The objective is that RWD curation should be reliable, with different abstractors producing the same results when performing the same task. This objective is not always satisfied, either due to remaining ambiguity in the policies and procedures, conflicting information in the chart, insufficient training, or human error.^6^

#### ML-extracted RWD

An ML model implements a function from inputs to outputs with parameters that are learned from data. Unlike a rules-based system that would require hard-coding of every possible nuance in clinical documentation, ML models can be trained to curate RWD using labeled data, such as the datapoints generated by human abstractors (Adamson, et al., in press, 2023). Whereas abstractors can be explicitly instructed on the policies and procedures, an ML model must learn those policies implicitly, based on statistical relationships it is exposed to during training. Its ability to correctly apply learned policies to unseen examples will depend on a number of factors, including the capacity of the model, the choice of feature representations for the inputs, and the scale, representativeness, and quality of the training data.

For many model architectures, the raw output of the function is a vector of continuous scores, even when the model is trained to perform a classification task. These scores provide additional information that can be useful for interpreting model outputs. In particular, the score corresponding to the predicted class can be interpreted as a measure of the model’s confidence. When a model is well-calibrated, low-confidence scores can be used to identify extracted datapoints that are potentially incorrect.

Importantly, the objective when applying ML for RWD curation is to extract explicit documentation of clinical information in the medical record, not to predict future outcomes or to infer latent attributes of a patient based on proxy measures. Therefore, model performance can be evaluated by comparing to human abstractors performing an equivalent task, and a high-performing model could be used as a “drop-in” replacement for or complement to manual curation.

#### Hybrid RWD

This work presents a method for using ML to scale the production of RWD even when model performance does not, on its own, match that of human abstractors. We refer to this method as “hybrid,” because it curates a single variable using both manual abstraction and ML extraction. The key aspect is that model confidence scores determine the source for each datapoint: curation is deferred to clinically-trained humans when model confidence falls below a threshold.

Our approach to curating a hybrid variable is as follows (Figure 1A). First, a full set of ML-extracted datapoints and confidence scores are obtained. The confidence scores are then compared to a variable-specific threshold. Datapoints with confidence scores below the threshold are sent to be manually abstracted. Finally, the results are merged into a single variable, with manually-abstracted datapoints replacing the low-confidence extracted datapoints.

**Figure 1:**
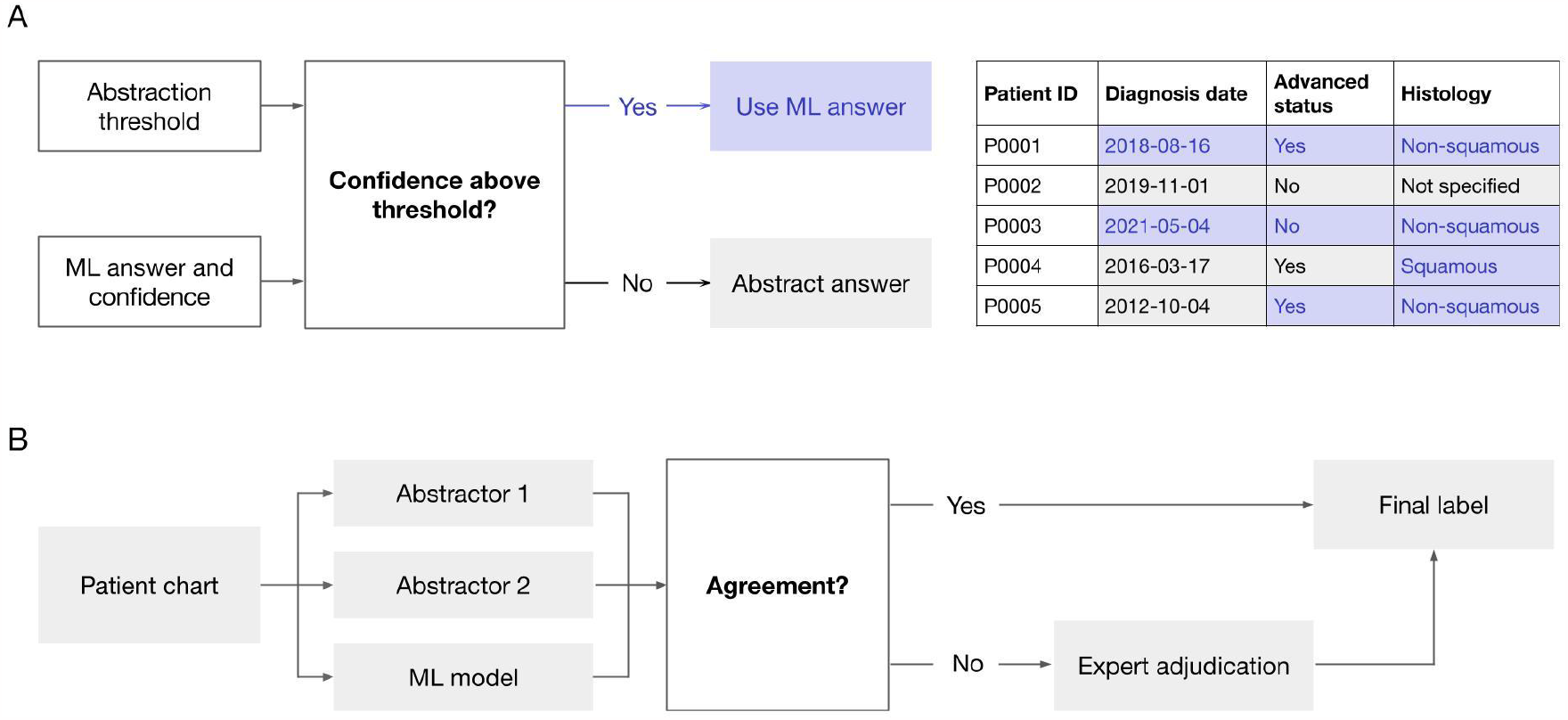
Diagrams illustrating the hybrid curation process. (A) The overall workflow for curating a hybrid variable. The confidence score associated with an ML datapoint is compared to a threshold, which determines whether the ML datapoint is used directly or if a human needs to manually abstract it. The resulting structured dataset includes a mixture of ML-extracted and manually-abstracted datapoints. (B) The label adjudication workflow. Labels used for selecting hybrid thresholds result either from a three-way agreement between model and abstractors or from the judgment of an expert adjudicator.

The threshold that determines whether a datapoint should be manually abstracted is a key parameter in this approach. With a lower threshold, less manual effort is required, but more model errors may be introduced. At the margin, increasing the threshold incurs additional manual effort to replace the lowest-confidence model outputs. The choice of threshold therefore governs a tradeoff between scale and quality. Yet this tradeoff may not always be straightforward: because human abstractors also make errors, overly stringent thresholds may yield diminishing or even negative quality returns. As a result, it is important to characterize empirically how effort and quality vary across thresholds.

#### Label adjudication

To evaluate hybrid variable performance at different thresholds, we curate a set of reference standard labels. Our approach aims to identify and remove label errors, which would otherwise distort our understanding of hybrid variable quality and cause us to overestimate the necessary amount of abstraction.^9^

The labels are curated in two stages (Figure 1B). Each chart is first reviewed independently by two human abstractors, and the resulting datapoints are compared to the model outputs. Any case with a disagreement among these three sources is then flagged for further review by an adjudicator. Adjudication is performed by clinical experts who have demonstrated consistently-high abstraction performance and been trained to conduct quality control on specific variables.

When reviewing disagreements, the adjudicators have access to the entire chart along with a record of the documents that abstractors accessed and any associated notes that they generated. They also have the option of consulting with a medical oncologist. Their goal is to identify the correct value for each datapoint as per the standardized policies and procedures, which may or may not be one of the values selected by the abstractors or model.

#### Threshold selection

Using the reference standard labels, we apply an empirical, simulation-based approach to threshold selection. A set of candidate thresholds is first identified by sampling between 0 and 1 in steps of 0.01. At each threshold, a simulated hybrid variable is formed. Where model confidence exceeds the threshold, the ML-extracted datapoint is used. Otherwise, we use the datapoint provided by the first abstractor during label curation. This mimics the process of replacing a low-confidence extracted value with the answer from a single abstractor. We then compute the error rate of the resulting hybrid variable and note the proportion of datapoints sourced from abstraction. Note that datasets may undergo subsequent cross-variable quality control checks that are not reflected in the error rate estimate.

After completing this process, we have a pair of metrics—abstraction rate and error rate—for each candidate threshold. These metrics quantify the tradeoff between effort and quality, allowing us to select an operating threshold that is appropriate for the expected use case. The following proof-of-concept study demonstrates that it is possible to find thresholds that produce satisfactorily low error rates with relatively small amounts of abstraction.

### Proof-of-concept methodology

#### Data sources

We used the nationwide de-identified Flatiron Health longitudinal EHR-derived database, comprising de-identified patient-level structured and unstructured data, curated via technology-enabled abstraction originating from ∼ 280 US cancer clinics (approximately 800 sites of care).^10,11^ Institutional Review Board approval of the study protocol was obtained prior to study conduct, and included a waiver of informed consent.

#### Cohort selection

For this proof-of-concept, we focused on a cohort of patients with non-small cell lung cancer (NSCLC). The cohort was initially limited to patients with a structured International Classification of Diseases (ICD) code for lung cancer and at least two EHR-documented clinic visits on or after January 1, 2011. We refined this cohort to patients with a clinically-documented NSCLC diagnosis as determined using ML model predictions^10^ and confirmation by two separate human abstractors. The analyses in this paper were performed on a random subset of this remaining cohort (N = 2373) that had been held out from the training of any ML models.

#### Variable definitions

This investigation considered three clinically meaningful real-world variables: the date of the patient’s initial NSCLC diagnosis, whether the cancer had ever been diagnosed as advanced, and the histology of the tumor.

##### Initial diagnosis date

The initial diagnosis date corresponds to the date on which the patient’s NSCLC diagnosis was confirmed. For the purposes of threshold selection in this investigation, the manually-abstracted or ML-predicted diagnosis date is considered “correct” if it is within 30 days of the reference standard date.

##### Advanced status

Advanced lung cancer is defined as stage IIIB, IIIC, IV (including IVA, IVB) at diagnosis or the subsequent development of recurrent or progressive disease for patients initially diagnosed with stage I–IIIA disease.

##### Histology

The histology variable has three possible classes: “Squamous cell carcinoma,” “Non-squamous cell carcinoma,” and “NSCLC histology, not otherwise specified” (NOS). Histologies without explicit mention of being squamous or non-squamous are grouped within the NOS class.

#### Manual abstraction

Manual abstraction of these variables followed a standardized set of policies and procedures defined in collaboration with oncologists to ensure that the operational definition matched the intended clinical concept. Abstraction was performed using Flatiron Health’s proprietary software platform. Within this system, unstructured documents were processed using Optical Character Recognition (OCR), categorized using a rules-based system, and surfaced to abstractors with supporting tools such as text search. Abstractors were required to have a minimum of 1 year of oncology experience, with more than 90% having one of the following licenses or certifications: nurse practitioner (NP), registered nurse (RN), licensed practical nurse (LPN), physician’s assistant (PA), doctor of pharmacy (PharmD), registered pharmacist (RPh), certified radiologic technologist (ARRT), certified clinical research associate (CCRA), certified clinical research professional (CCRP), or certified clinical research coordinator (CCRC). All abstractors passed an abstraction assessment pre-hire along with task-specific training and knowledge assessments. The same basic qualifications applied to adjudicators, who were also trained on quality control procedures.

#### ML model construction

The ML models first scan charts for mentions of clinically-relevant terms and extract “snippets” of text surrounding these mentions. The snippets are then vectorized and passed into a classifier architecture.^10,12^

For the initial diagnosis date and advanced status variables, the model is a deep neural network based on dense word embeddings and a bidirectional GRU encoder^13^ Diagnosis date predictions are made for multiple timepoints and then aggregated to identify the most likely date. For the histology variable, the classifier uses TF-IDF vectorization and gradient-boosted decision trees.^14^

The three models were trained independently on overlapping but distinct sets of patient charts. Training sets varied by model based on availability of labels but included more than 25,000 examples in all cases. Training was performed using a supervised objective with manually-abstracted datapoints serving as labels. All models were constructed with a final softmax layer and trained with a cross-entropy loss function to encourage good calibration. Note that the hybrid methodology generalizes across these distinct architectures with no further accommodations.

#### RWD-defined cohort comparisons

Given the emergent role of ML-extracted RWD for scientific and health outcomes research,^15^ it is critical to go beyond accuracy metrics and establish that hybrid variables are fit for purpose when used in retrospective research. As such, we replicated a cohort selection task using either a fully-abstracted or hybrid version of the NSCLC dataset and compared the resulting patient characteristics. The goal of this analysis was to evaluate whether the selected cohorts remained similar despite differences in the underlying data curation methodology.

Cohort selection included patients who had an initial diagnosis date on or after 2018, advanced NSCLC, and non-squamous histology. We selected these criteria to identify patients who might benefit from recent advances in NSCLC management, including molecular testing and targeted therapies. For the two resulting cohorts, we compared baseline patient characteristics available in structured EHR fields, using categorical variables to define practice type, region, sex, race, age at first visit, year at first visit, and follow-up duration from first visit. Patient characteristic distributions were summarized with counts and proportions, and cohort similarity was evaluated using the Absolute Standardized Mean Difference (ASMD) metric. We report the point estimate of ASMD as well as bootstrap confidence intervals generated with 100 resamples of the selected cohorts, using the conventional cutoff of ASMD less than 0.1 to indicate “cohort similarity.”^16^

## Results

### Overall performance metrics

We evaluated the performance of single-abstracted and ML-extracted data using the reference standard labels, which provide a common basis for comparison. The diagnosis date variable was evaluated using a 30-day agreement metric, while the advanced status and histology variables were evaluated using weighted-macro-average F1 over each class.

Overall performance for both single-abstracted and ML-extracted data exceeded 0.9 across all three variables (Table 1). When compared directly, manual abstraction significantly outperformed the ML model, although the difference was small in absolute terms (< 0.05 pts). Therefore, replacing all abstracted datapoints with ML datapoints would incur a small but measurable decrease in the quality of the dataset.

**Table 1:**
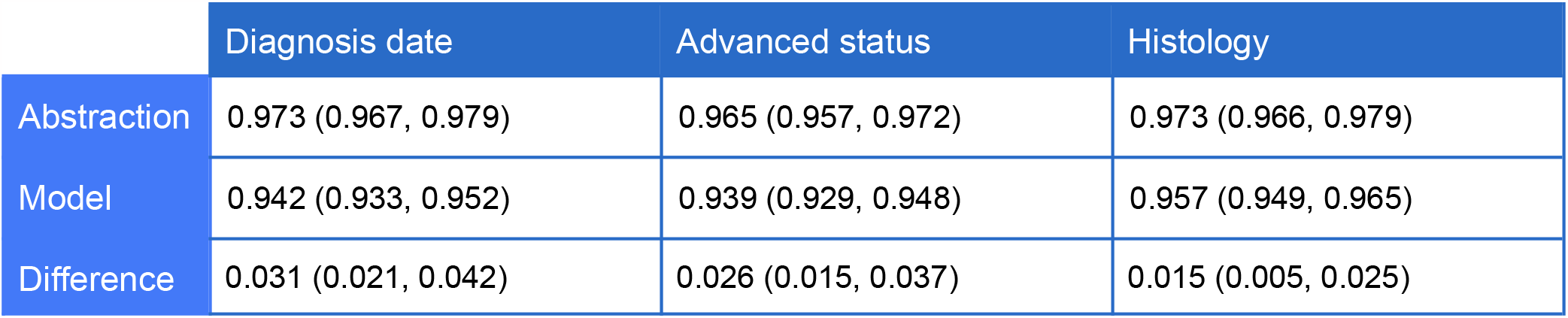
Performance metrics when compared to the reference standard. Diagnosis date scores represent accuracy within a 30-day window. Advanced status and histology scores represent weighted macro-average F1. 95% confidence intervals were calculated using a bootstrap.

|  | Diagnosis date | Advanced status | Histology |
| --- | --- | --- | --- |
| Abstraction | 0.973 (0.967, 0.979) | 0.965 (0.957, 0.972) | 0.973 (0.966, 0.979) |
| Model | 0.942 (0.933, 0.952) | 0.939 (0.929, 0.948) | 0.957 (0.949, 0.965) |
| Difference | 0.031 (0.021, 0.042) | 0.026 (0.015, 0.037) | 0.015 (0.005, 0.025) |

### Model confidence scores

Overall, model confidence was high (Figure 2). Median confidence scores were 0.95 for diagnosis date and advanced status and 0.96 for histology; at least two thirds of the extracted datapoints had a confidence score above 0.9 (diagnosis date, 67%; advanced status, 69%; histology, 80%).

**Figure 2:**
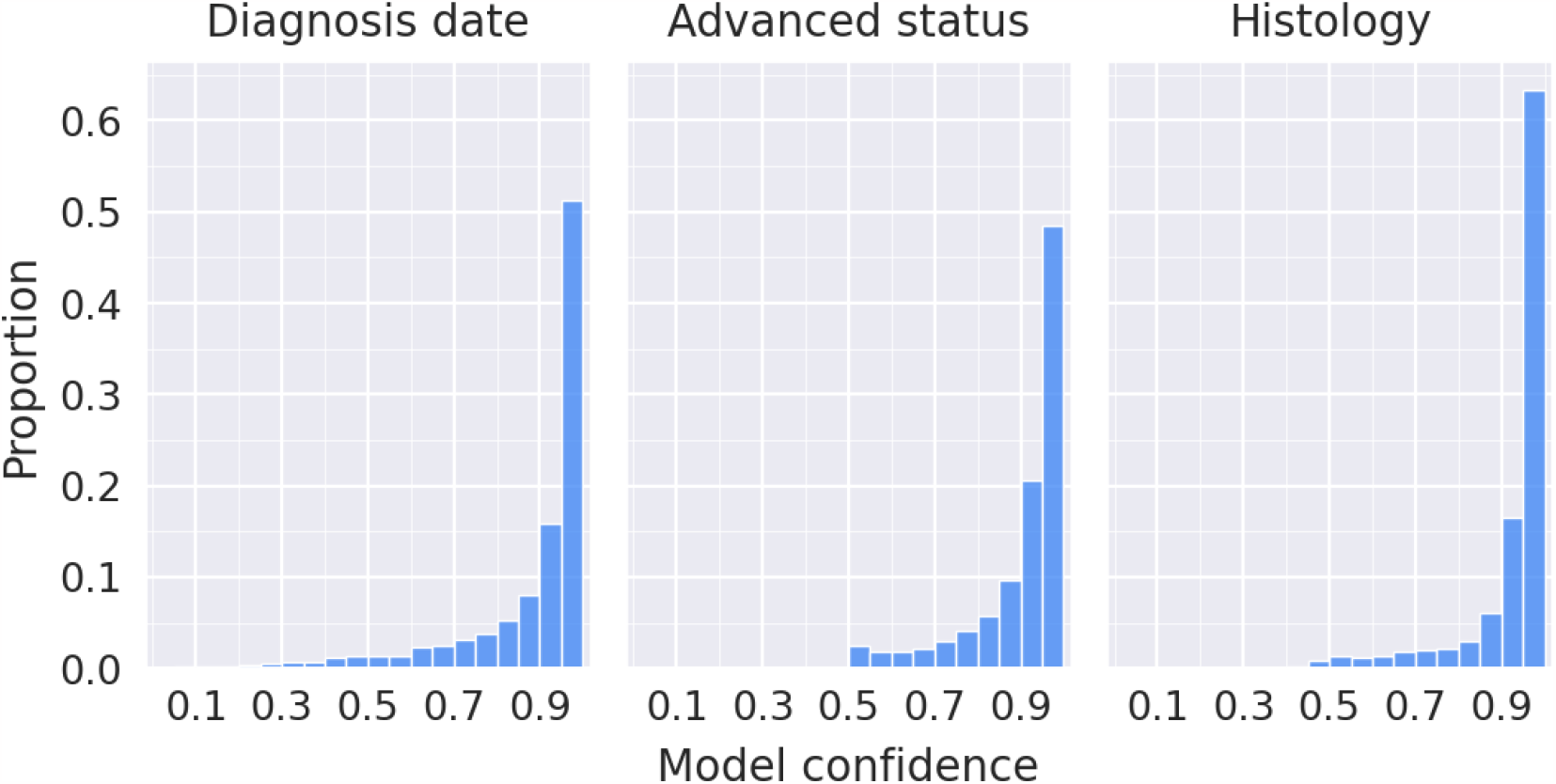
Distributions of model confidence for three variables of interest. Each bin width is 0.05.

For the hybrid methodology to be effective, most model errors should occur when there is low confidence in the extracted datapoint. Consistent with this requirement, more than 50% of all model errors occurred in the lowest decile of model confidence. Abstraction errors were also more likely to occur when the model’s confidence was low, but the abstraction error rate in the lowest decile was less than half the model error rate (Figure 3). Beyond the lowest two deciles of confidence, model error rates appeared equivalent to or lower than abstractor error rates.

**Figure 3:**
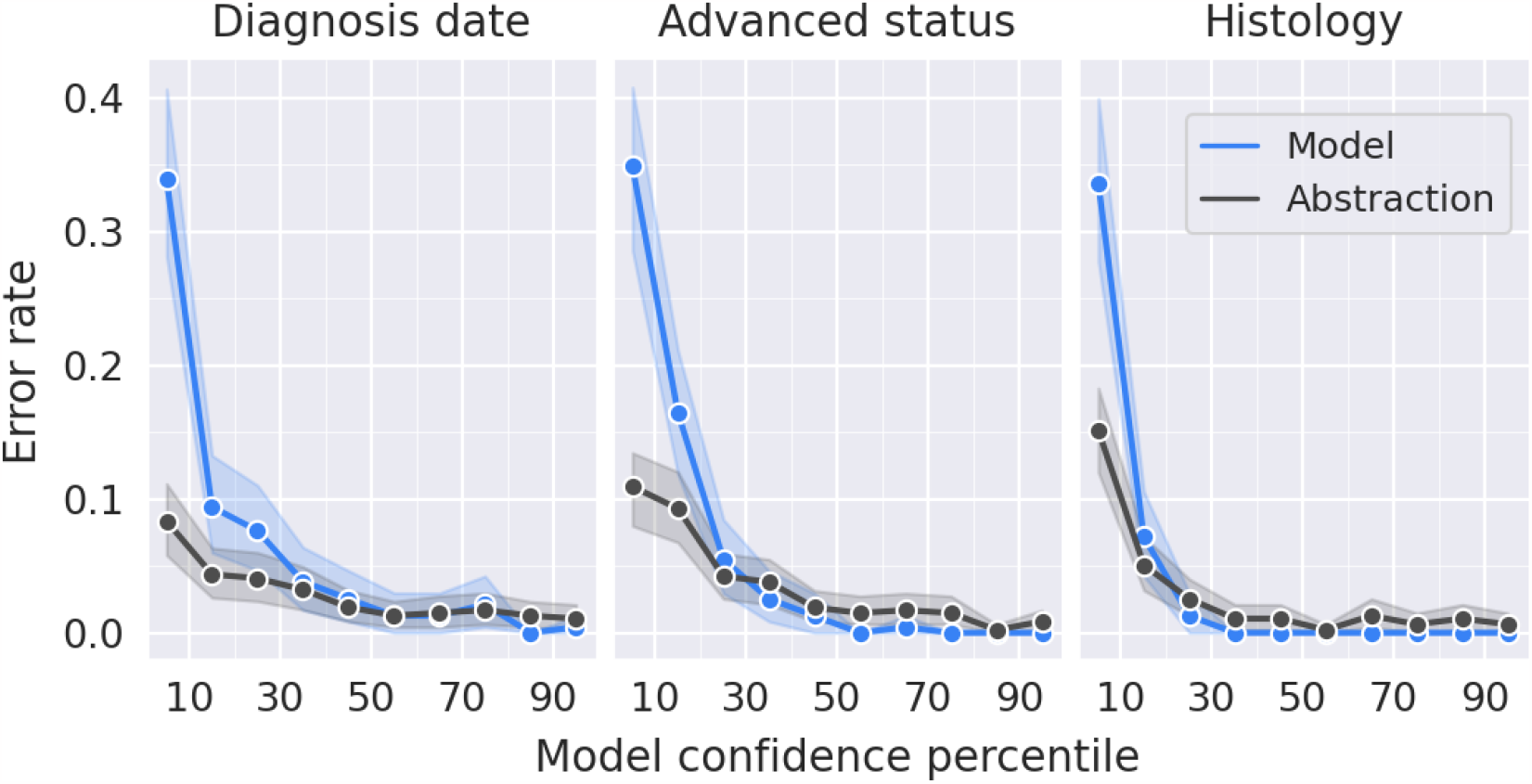
Error rates for extracted and abstracted data when stratified by deciles of model confidence. Error bands represent 95% bootstrap confidence intervals.

### Hybrid variable evaluation

We simulated hybrid variables across a range of candidate thresholds and evaluated them using the reference standard labels. At each candidate threshold, we obtained two metrics: the proportion of the cohort that would be deferred to manual abstraction and the error rate of the resulting hybrid variable (Figure 4). Consistent with our expectation of a tradeoff between abstraction effort and data quality, the highest error rates were observed at thresholds resulting in 0% abstraction. Raising the threshold slightly to include some abstracted datapoints rapidly decreased error rates: the absolute difference between hybrid and fully abstracted variables fell below 1% when abstracting no more than 10% of the cohort for any variable (diagnosis date, 9.9%; advanced status, 5.4%; histology, 0.8%).

**Figure 4:**
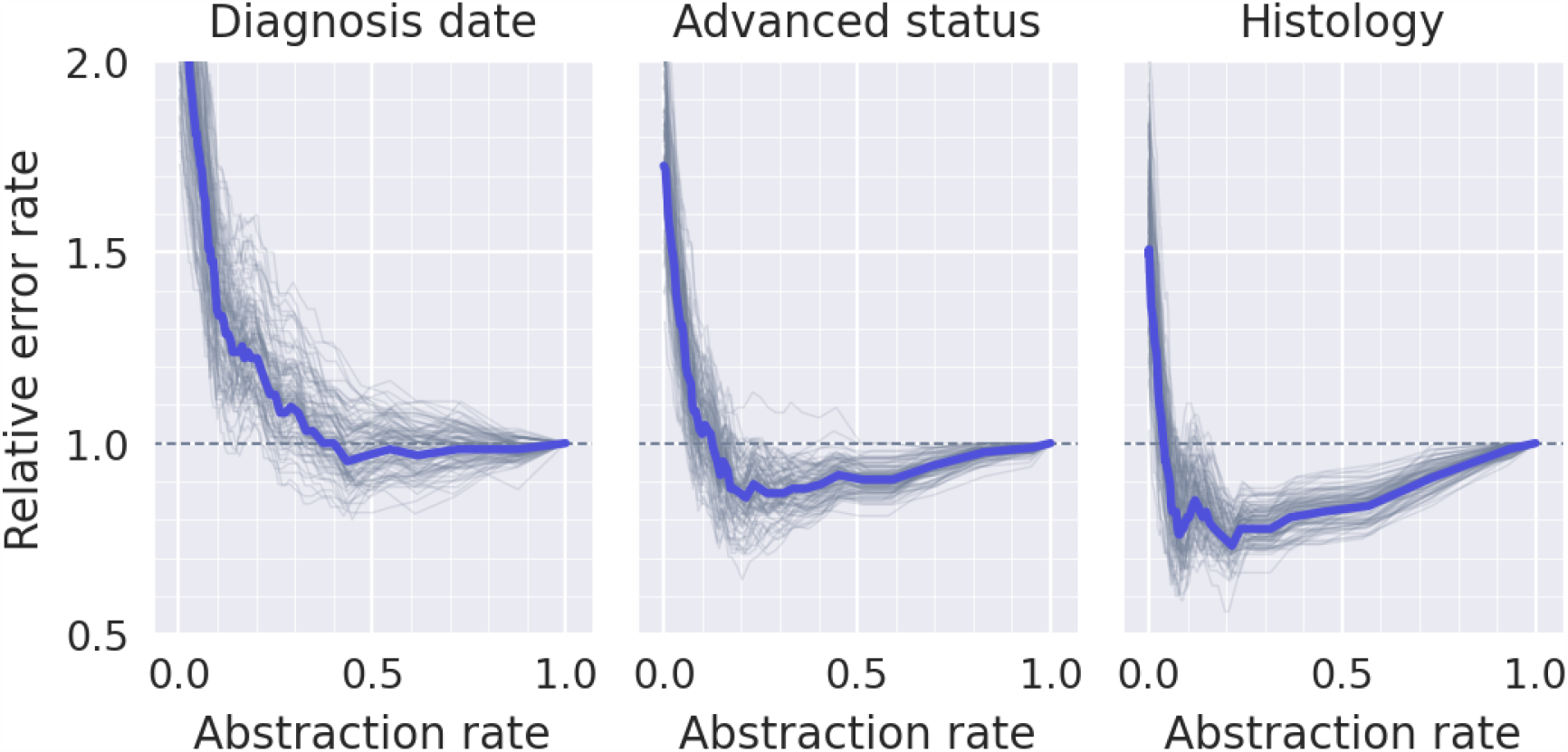
Hybrid variable evaluation. Curves are drawn by simulating performance across a range of candidate thresholds (evenly spaced by 0.01). The x axis shows the abstraction rate (proportion of the cohort with a confidence score below the threshold), and the y axis shows the corresponding hybrid variable error rate. Error rates are normalized by the value for a fully-abstracted variable (abstraction rate = 1); relative error rates below 1 indicate fewer errors in the hybrid variable. Thick blue lines represent a curve derived from the entire cohort; thin gray lines represent curves derived from bootstrap replicates of the cohort (100 total).

Surprisingly, the lowest error rates were not achieved with thresholds resulting in 100% abstraction. When the model was highly confident, it tended to be more accurate than human abstractors (Figure 3). Therefore, the error-minimizing threshold for both advanced status and histology required abstracting less than a quarter of the cohort (advanced status, 21.2%; histology; 21.5%). At these thresholds, hybrid error rates were significantly lower than full-abstraction error rates (*P* < 0.05; bootstrap test). In contrast, the diagnosis date error rate was numerically lowest when abstracting a minority of the cohort (43.5%), but it was not significantly lower than the full abstraction error rate at any threshold. Therefore, the hybrid methodology allows us to increase cohort sizes while maintaining—and in some cases improving—the accuracy of the curation process.

### RWD-defined cohort comparisons

While there are several potential objectives for selecting a confidence threshold, for this investigation we aimed to minimize abstraction effort without increasing error rates relative to manual abstraction (i.e., we chose a threshold corresponding to the left-most intersection between the blue and dashed lines in Figure 4). This approach resulted in an abstraction workload of 37.8%, 13.0%, and 4.0%, respectively, for the diagnosis date, advanced status, and histology variables. A total of N=517 patients were selected using abstracted variables, while N=526 patients were selected using hybrid variables. There were N=495 (90%) patients in the intersection of the cohorts. Distributions of patient characteristics were consistently similar between cohorts, with ASMDs ranging from 0.012 to 0.034 (Figure 5). All patient characteristics had ASMDs less than 0.1. Therefore, cohorts curated using hybrid methodology are similar to cohorts derived using only abstraction, even when using automated extraction for the majority of cases.

**Figure 5:**
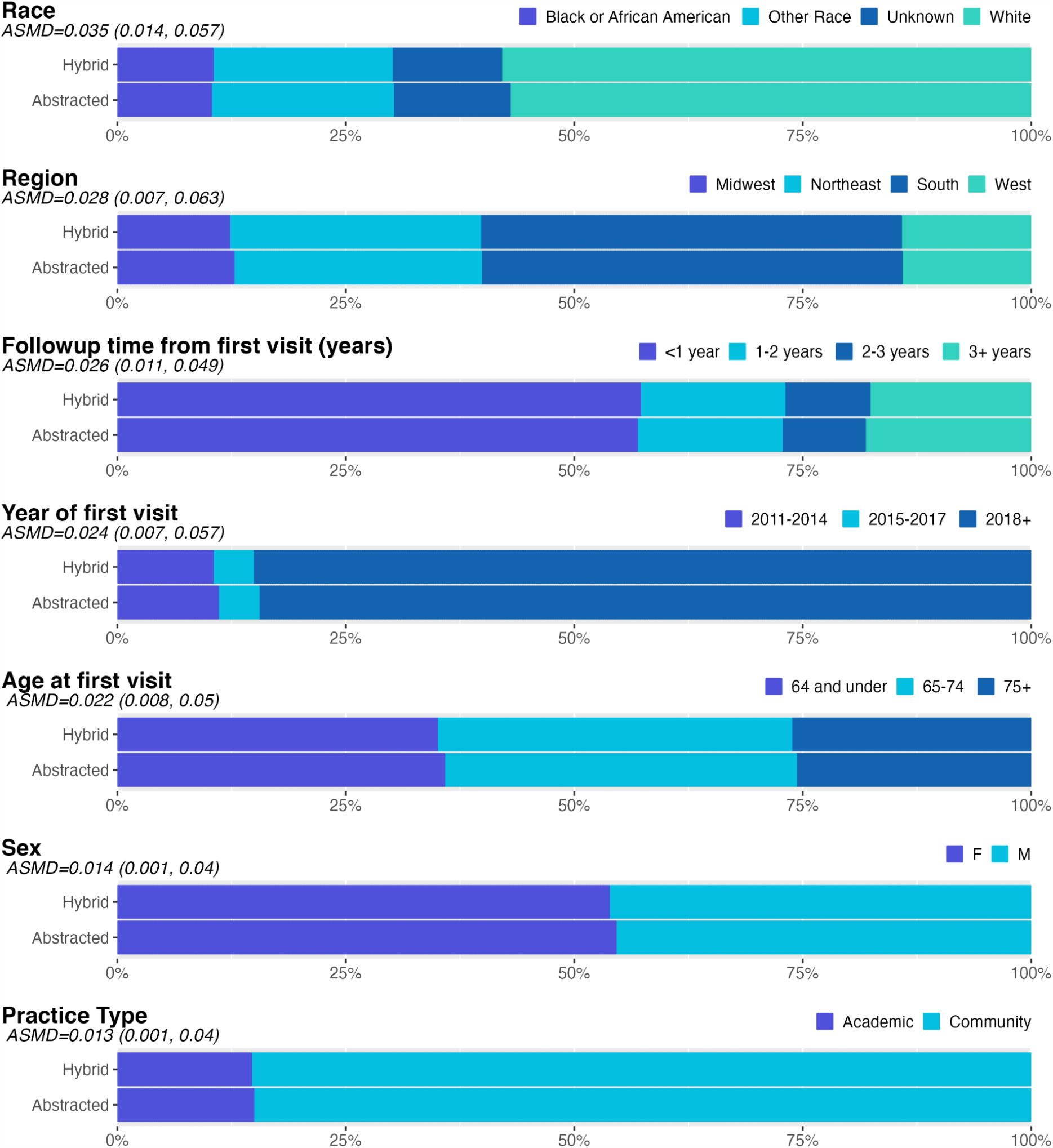
Comparison of cohorts selected using hybrid variables (N=526) vs abstracted variables (N=517). Stacked bar plots show the proportions of categories for each patient characteristic. Cohort selection included patients who had an initial diagnosis date on or after 2018, advanced NSCLC, and non-squamous histology. ASMD: Absolute Standardized Mean Difference is calculated for each characteristic comparing hybrid to abstracted cohorts (95% confidence intervals derived using bootstrap resampling). ASMD < 0.1 is the typical cutoff to define “similarity.”

## Discussion

We introduce a hybrid curation methodology that integrates clinical expertise and machine learning to scale the production of high-quality real-world data. By relying on models when confident, the methodology focuses abstraction effort on the hardest tasks. This increases the achievable cohort size for a given quantity of abstraction resources. As an example, we demonstrate the feasibility of abstraction rates that imply a nearly three-fold expansion of cohort sizes with no additional human effort.

Critically, we show that scaling need not come at the expense of quality. Given the complexity of information in an EHR, abstraction remains important: humans can synthesize disparate data sources, incorporate contextual information, and reason about edge cases. Nevertheless, we find that ML can outperform abstraction in some situations. Why might this be? ML excels at surfacing “needles in the haystack” of voluminous medical records. And even highly-trained abstractors suffer from cognitive fatigue and make mistakes when performing repetitive tasks. Hybrid data curation allows us to leverage the strengths and minimize the weaknesses of each method.

Unstructured data has long been recognized as a critical source of information about patient treatment and outcomes, particularly in oncology.^17^ Yet scalable extraction remains a challenge.^5^ Automated methods and human abstraction are generally viewed as competing alternatives.^18,19^ This work offers a new paradigm in which ML and human abstraction are jointly leveraged.

Because human abstraction is imperfect, label adjudication is key to the hybrid curation process. Consider the following: were single-abstracted data treated as the reference standard, a hybrid system could succeed only if the model agreed with every abstracted datapoint, even when incorrect. By conducting a three-way comparison and adjudicating disagreements, we establish a robust evaluation framework that overcomes these limitations. This allows us to more objectively assess performance and optimize the balance between manual abstraction and automated extraction. While other authors have proposed adjudication frameworks for labeled data, these generally rely only on pairwise comparisons.^20 21^

Advantages of our thresholding methodology include its generalizability across model architectures—only requiring confidence scores to be rank calibrated—and the flexibility it affords for balancing scale and quality as appropriate for a given use case. One could choose threshold values that maintain quality relative to abstraction or trade either objective against the other as needed. While the models used here are highly accurate, we expect that the method can still be beneficial when there is a larger gap between abstractors and models, although the amount of abstraction required to maintain quality may be higher. Notably, our framework incorporates the special cases of using only abstraction (threshold of 1) or only ML-extraction (threshold of 0), and it would reveal when one of these choices is optimal.

This work represents a natural step in the broader effort to combine human abstraction with ML for scalable data curation. Previously, ML has been employed to automate cohort identification upstream of abstraction, filtering out patients that are unlikely to meet cohort inclusion criteria.^10^ Likewise, Marucci-Wellman et al.^21^ used ML to identify and filter out “easy” abstraction tasks, although in the absence of reference standard labels, they guide thresholds solely by resource constraints and do not consider quality objectives. Alternatively, human expertise can directly improve model performance through a feedback mechanism, where the model selectively queries experts to provide additional input on “hard” cases.^22^ This iterative approach has proven useful for reducing the effort required to extract diagnosis dates from unstructured records, akin to one of the tasks here.^23^ Other authors have proposed schemes to allocate cases between humans and models,^24,25^ although they primarily focus on the model training process.

An alternate approach for increasing real-world cohort sizes is to link and pool observations across multiple data sources, such as structured EHR fields, administrative claims, and tumor registries.^26,27^ An advantage of our method is that both abstracted and ML datapoints are based on a common source, reducing heterogeneity and potential bias. As illustrated in the cohort comparison, data curation via our method produced very similar cohorts compared to abstraction alone. Such scalable, high-quality extraction from unstructured text data facilitates rapid and accurate production of real-world evidence.^28^

## Conclusion

This work offers a novel approach to scalable data curation that jointly leverages ML and human expertise. The proposed hybrid framework is quite flexible to different data types and ML models, making it generally applicable to a variety of curation tasks. Hybrid curation holds significant potential for real-world evidence, enabling the study of larger patient cohorts while remaining fit-for-use in a range of research and policy applications.

## Data Availability

The data that support the findings of this study have been originated by Flatiron Health, Inc. Requests for data sharing by license or by permission for the specific purpose of replicating results in this manuscript can be submitted to.

